# COVID-19 peak estimation and effect of nationwide lockdown in India

**DOI:** 10.1101/2020.05.09.20095919

**Authors:** R V Belfin, Piotr Bródka, B L Radhakrishnan, V Rejula

## Abstract

There was a fury of the pandemic because of novel coronavirus (2019-nCoV/SARS-CoV-2) that happened in Wuhan, Hubei province, in China in December 2019. Since then, many model predictions on the COVID-19 pandemic in Wuhan and other parts of China have been reported. The first incident of coronavirus disease 2019 (COVID-19) in India was reported on 30 January 2020, which was a student from Wuhan. The number of reported cases has started to increase day by day after 30 February 2020. The purpose of this investigation is to provide a prediction of the epidemic peak for COVID-19 in India by utilizing real-time data from 30 February to 14 April 2020. We apply the well-known epidemic compartmental model “SEIR” to predict the epidemic peak of COVID-19, India. Since we do not have the complete detail of the infective population, using the available infected population data, we identify the R_0_ by using polynomial regression. By using the third-order polynomial equation, we estimate that the basic reproduction number for the epidemic in India is R_0_ = 3.3 (95%CI, 3.1–3.5), and the epidemic peak could be reached by September 2020.

## 1. Introduction

During December 2019, a new virus named CORONA (1), later renamed COVID-19, devised an outbreak in Wuhan, a city in the Hubei region, China. Subsequently, COVID-19 started to spread to the other parts of China and many other countries around the globe. By the end of January 2020, there was a global COVID-19 positive case of 9,776 individuals, and the number of death casualties crossed 213. The WHO announced it as a worldwide public health crisis (2). COVID-19 worldwide death causalities rose to 811 by 9th February and surpassed the total death casualties of the SARS epidemic during 2003. Afterward, the number of COVID-19 positive cases started to grow at a rapid pace(3). As China and other COVID-19 affected nations reacted quickly to this pandemic, it is crucial to understand the severity, the total number of infected people, COVID confirmed cases, total death, basic reproduction number, and other significant determinants related to this deadly pandemic. Before-mentioned information may help the health care administrators in the respected regions and the city health care executives to make bold and informed choices (4).

As of 5th May, globally, COVID affected cases rose to 3,646,225, and the death toll rose to 252,941 individuals; meanwhile, on a positive note, recovered count increased to 1,206,276 people (5). Countries such as the USA, Spain, Italy, Germany, and the United Kingdom have the highest number of COVID-19 cases, respectively(6). COVID-19 has spread to 210 countries and territories around the globe, and two international conveyances named Diamond Princess cruise ship harbored in Yokohama, Japan, and Holland America’s MS Zaandam cruise ship(7).

In India, as of 29th April 2020, according to the official website <u>of</u> India, COVID-19 affected 22629 people, recovered 7695 people, and a death toll of 1007 individuals, respectively (8). India reported its first case (9) on 30th January 2020 in a coastal state named Kerala in the southern part of India, which was a student who traveled from Wuhan, China. Subsequently, the COVID-19 positive cases started growing day by day because of many other travelers and tourists arrived through airways (10). The detail of the week by week increase in COVID-19 cases has been given in table 1.

**Table 1.** COVID-19 weekly reported and accumulated cases in India.

| Week | Number of Newly Reported Cases | Number of Accumulated Cases |
| --- | --- | --- |
| 30 January – 1 February | 1 | 1 |
| 2 February – 8 February | 2 | 3 |
| 9 February – 15 February | 0 | 3 |
| 16 February – 22 February | 0 | 3 |
| 23 February – 29 February | 0 | 3 |
| 1 March – 7 March | 31 | 34 |
| 8 March – 14 March | 48 | 82 |
| 15 March – 21 March | 332 | 414 |
| 22 March – 28 March | 987 | 1,401 |
| 29 March – 4 April | 3,588 | 4,989 |
| 5 April – 11 April | 8,446 | 13,435 |
| 12 April – 18 April | 16,365 | 29,880 |
| 19 April – 27 April | 29,451 | 59,251 |

According to the number of cases in table 1, there is a significant spread of COVID-19 that happened in India. The most significant concern of the Indian public is the duration of the pandemic that is extending at the time of poorly shaped economy. (Mathew, 2019; Pesek, 2020). The purpose of the study is to predict the pandemic peak in India and assists the public and the government in taking predictive measures to keep the safeguard Indian people in this worst pandemic situation.

The numbers presented in table 1 have uncertainty between reported cases and accumulated cases. The reported cases have grown exponentially since the affected person may spread the infection to its fellow beings (4,13,14). The accumulated COVID-19 cases are high because of the unavailability of drugs to treat the disease and an insufficient number of test kits to detect it (15). Recently India procured some rapid COVID-19 test kits from China, and most of the kits showed inaccurate results (16). This study takes this uncertainty in the infected and the accumulated population count and applies a mathematical model for prediction. More specifically, the study assumes that diagnostics can identify only p(0<p≤1) fraction of infected Indian citizens. The value p is the probability of an infected Indian from the whole population. Also, For Analysis, we have assumed the first date of infection in India as of March 1, 2020, because after January 30^th^ and February 1^st^ there were no COVID-19 infected cases reported in India till February 30^th^.

## 2. Related Work

Wilder-Smith (3) modeled the COVID-19 outbreak of Diamond Princess ship using the SEIR model. After calibrating the model with transient functions, the results show that population density creates high risk. The contact rate and R_0_ are dependent on the density of the population. Containment measures reduced the initial R_0_ 14.8 to 1.78. Kuniya (17) predicts the COVID-19 peak of Japan using the SEIR model. Infection rate β was estimated by using the best fit procedure least square with poison noise. The results show that R_0_ = 2.6 (95%CI, 2.4−2.8). Interventions delay the epidemic peak delayed the peak from 179 days to 243; it reduces the size of the epidemic. Ranjan (18) estimated the peak prediction of India using the SIR model. The results show the R_0_ = 1.4-3.9. Early lockdown in India aid in reducing the spread of COVID-19 to a considerable number. Prem (19) studied the effect of countermeasures in China using the SEIR model. The authors made a contact matric for distinct intervention scenarios to study the impact of social distancing. The results show that social distancing was a valid measure, and early removal of social restrictions would create another peak. Wang (20) estimated the epidemic trend in China using the SEIR model. The results show that R_0_ was 1.9, 2.6, and 3.1 before implementing any countermeasures. After applying the countermeasures in phases, simultaneously, R_0_ also reduced in each phase ranging from 3.1 to 0.5.

Based on the review of different prediction models, we realized that the interventions are useful to delay the outburst while predicting the influencing variables should be incorporated to get a better result. However, prediction techniques assist the countries in formulating a better containment plan. The containment plan may differ based on the preparedness status of each country and the response plan (4).

## 3. Methods

In this section, we present the basic concepts of the SEIR compartmental epidemic model, the basic concepts related to the SEIR model, and definitions that we have used.

### 3.1. Model

This study applies the well-known SEIR epidemic model for prediction. The state diagram of the Susceptible-Exposed-Infected-Removed model depicted in Fig 1. SEIR will be an appropriate model for predicting COVID-19 because it has a considerable post-infection incubation period in which the exposed person is not infectious.

**Fig 1:**
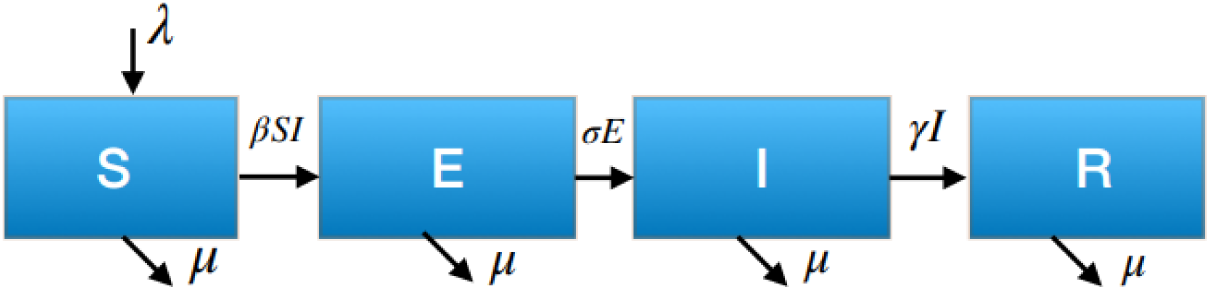
State Diagram for the SEIR epidemic model.

In Fig 1, the notations S, E, I, R denotes the compartments in the model named Susceptible, Exposed, Infected, and Removed, respectively. β is the effective transmission rate, λ is the “birth” rate of susceptible population, μ is the mortality rate, σ is the onset rate from latent to infected and γ is the removal rate (recovery or death). For this prediction, the birth rate λ and the mortality rate μ are insignificant; hence we define the SEIR model with our new birth and mortality in the eq (1).

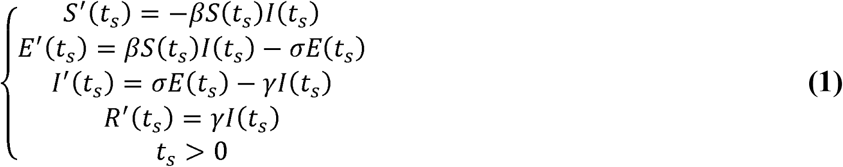

In equation (1), S(t_s_), E(t_s_), I(t_s_), R(t_s_) denotes the Susceptible, Exposed, Infectious and Recovered population in time slice t_s_, respectively. Similarly, 1/σ, 1/γ is the mean incubation period and mean transmittable period, respectively. This study assumes the per unit time as one day.

By using the numbers from the previous studies given in Table 2, the mean incubation 1/σ has been fixed as 1/σ = 5; thus, σ = 0.2 and recovery rate γ = 0.1, respectively. Also, S + E + I + R has been fixed to be one so that ultimately each population will be equated with the proportion to the total population.

**Table 2.**
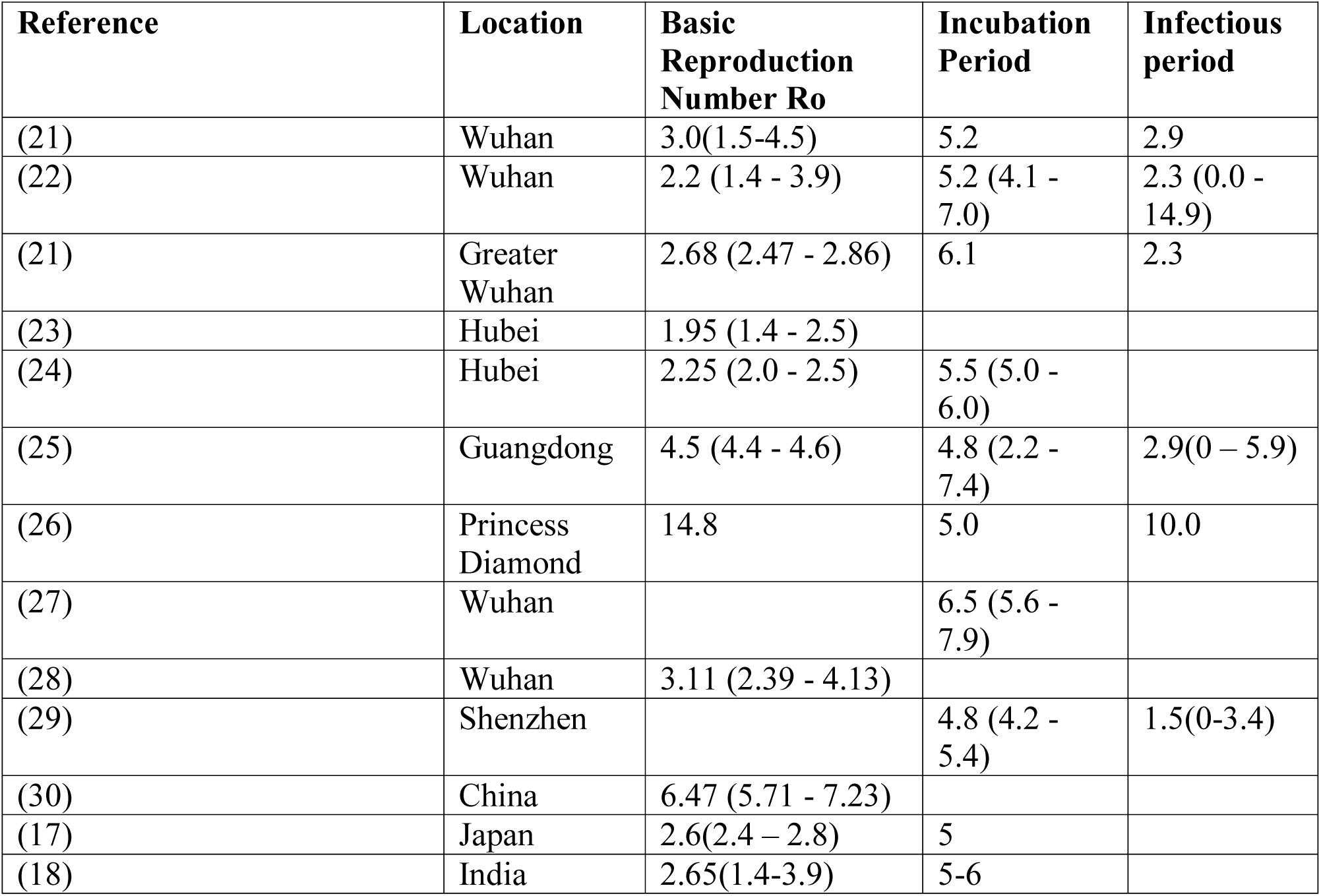
Estimations of various parameters for epidemic models from previous research.

| Reference | Location | Basic<br>Reproduction<br>Number $R_0$ | Incubation<br>Period | Infectious<br>period |
| --- | --- | --- | --- | --- |
| (21) | Wuhan | 3.0(1.5-4.5) | 5.2 | 2.9 |
| (22) | Wuhan | 2.2 (1.4 - 3.9) | 5.2 (4.1 - 7.0) | 2.3 (0.0 - 14.9) |
| (21) | Greater<br>Wuhan | 2.68 (2.47 - 2.86) | 6.1 | 2.3 |
| (23) | Hubei | 1.95 (1.4 - 2.5) |  |  |
| (24) | Hubei | 2.25 (2.0 - 2.5) | 5.5 (5.0 - 6.0) |  |
| (25) | Guangdong | 4.5 (4.4 - 4.6) | 4.8 (2.2 - 7.4) | 2.9(0 - 5.9) |
| (26) | Princess<br>Diamond | 14.8 | 5.0 | 10.0 |
| (27) | Wuhan |  | 6.5 (5.6 - 7.9) |  |
| (28) | Wuhan | 3.11 (2.39 - 4.13) |  |  |
| (29) | Shenzhen |  | 4.8 (4.2 - 5.4) | 1.5(0-3.4) |
| (30) | China | 6.47 (5.71 - 7.23) |  |  |
| (17) | Japan | 2.6(2.4 - 2.8) | 5 |  |
| (18) | India | 2.65(1.4-3.9) | 5-6 |  |

Considering the population in India, the study assumes that one infective individual identified at time slice t=0 among total N=1,350,000,000 number of people in India (31). This implies γ (0) = pI(0) N = 1, Where

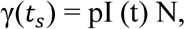

denoted the number of infectious individuals identified at time slice t_s_. Thus, we obtain I(0) = 1/(pN). The study assumes that there are no exposed and removed people during the time slice t_s_=0, which implies E(0) = R(0) = 0, and hence,

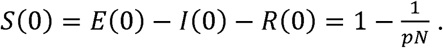

Based on the previous works (17), we assume the p-value rages from 0.01 to 0.1. The basic reproduction number R^0^ denotes the secondary spread made by one infected individual(32), is calculated as the maximum eigenvalue of the next generation matrix FV^-1^ (33,34), where

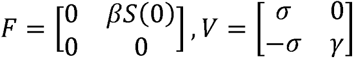

Therefore we obtain,

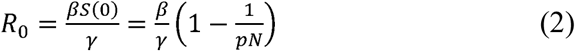

### 3.2. The sensitivity of the Basic Reproduction Number

The basic reproduction number R_0_ is independent of the onset rate σ. The sensitivity of R_0_ to other parameters β, γ, p are calculated as:

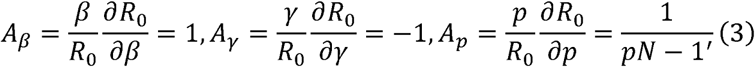

Where, *A_β_*, *A_γ_*, *A_p_* denotes the normalized sensitive indexes for β, γ, p, respectively. As we note from the equation (3) that the k time’s increase in β (respectively γ) results in the k (respectively k^-1^) time’s increase in R_0_. Most important, we note from equation (3) that A_p_ ≈ 0 if p ≥ 1.0 x 10^−7^. This implies that the identification rate p in a realistic range almost does not affect the size of R_0_.

### 3.3. Estimation of the infection rate

Let y(t_s_), t_s_ = 0,1,…,45 be the number of daily reported cases of COVID-19 cases in India from 1^st^ March (t_s_ =0) to 14^th^ April 2020 (t_s_ =45). We perform a polynomial regression to predict the infection rate β of COVID-19 in India.

#### Description 1

Polynomial Regression is a regression algorithm that generations the relationship within a dependent (y) and independent variable (x) as n^th^ degree polynomial. The Polynomial Regression equation is given below:

As given in the equation, in Polynomial regression, the standard features of the variables are transformed into the Polynomial features of necessary degree (2, 3, .., n) and then modeled by a linear model.” As we see in Fig 2, the data points are not linear, so we need to use a non-linear method to predict this rate of infection *β*.

**Fig 2:**
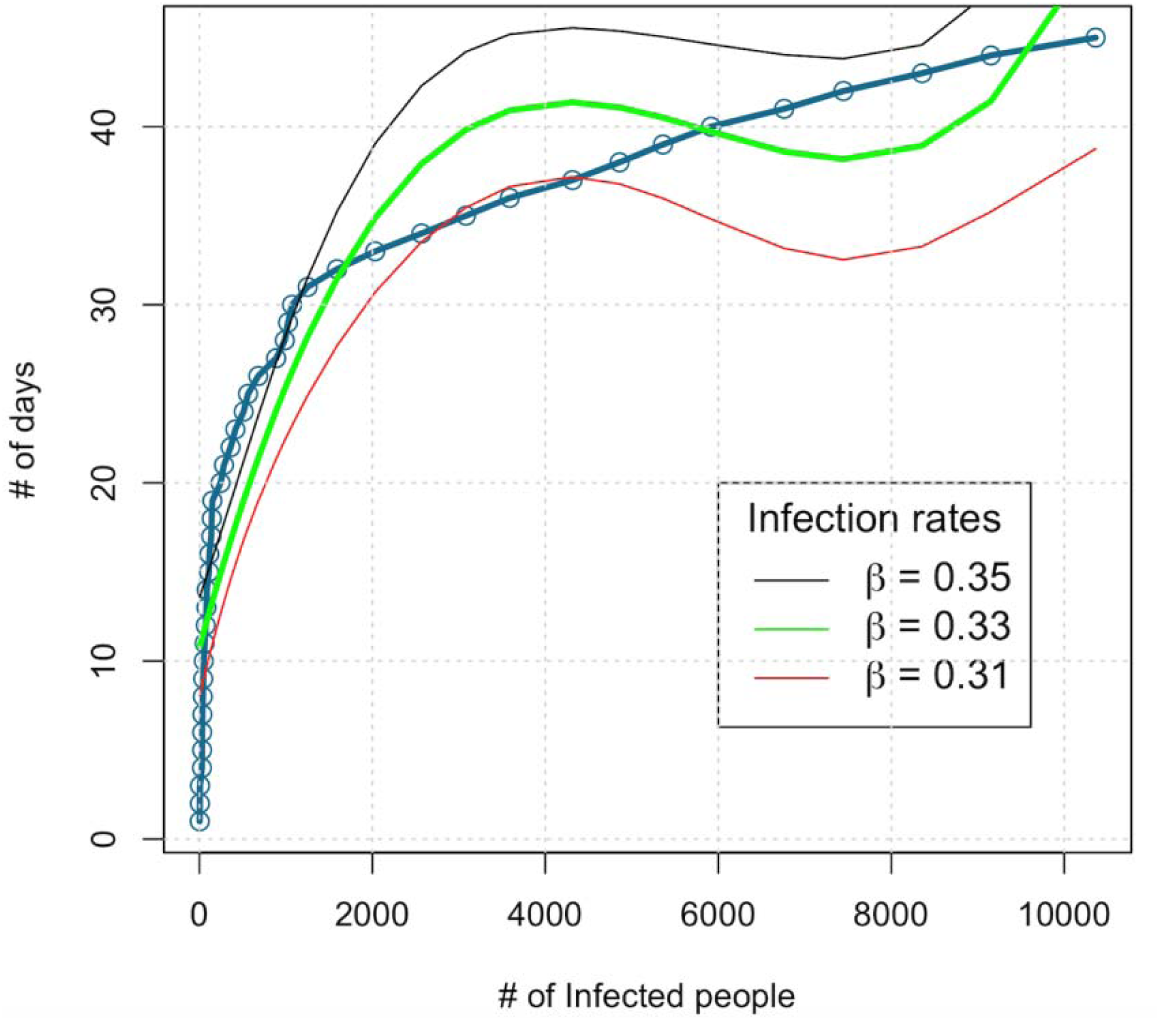
The comparison of Y(t_s_) with the estimated infection rate β and the number of daily reported cases of COVID-19 in India from 1st March (t_s_ = 0) to 14th April (t_s_ = 45)

**Fig 2.**
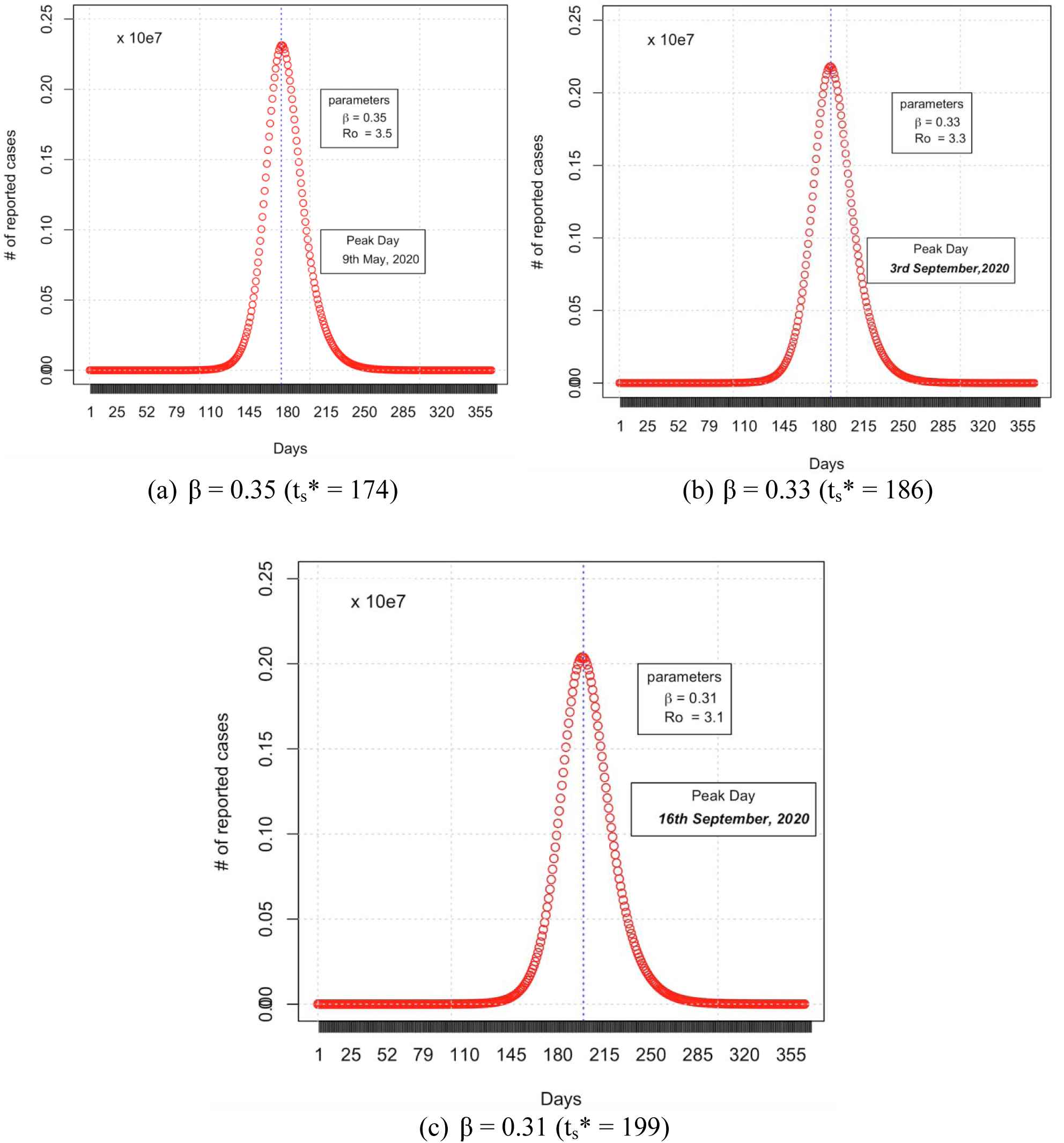
Time variation of the number Y(t) of the infective individuals who are identified at the time slice t_s_ (0< t_s_ <365) for p=0.1. The dotted lines represent the epidemic peak t_s_*.

Thus we obtain β estimated as 0.33 (95% CI, 0.31-0.35). The polynomial regression fit has been given in Fig 2. Then, we obtain the value or R_0_ from the equation (2). We get an estimation of R_0_ as 3.3 (95%CI, 3.1-3.5). The values identified and used for the COVID India Peak prediction has been given in Fig 2.

**Table 3:** Parameter values for predicting COVID-19 India.

| Parameter | Description | Value | Reference |
| --- | --- | --- | --- |
| $\beta$ | Infection Rate | 0.33(95%CI, 0.31-0.35) | Estimated |
| $R_0$ | Basic Reproduction Number | 3.3(95%CI, 3.1-3.5) | Estimated |
| $\sigma$ | Onset Rate | 0.2 | (21) |
| $\gamma$ | Removal Rate | 0.1 | (35) |
| $N$ | Total population in India | $1.35 \times 10^9$ | (31) |
| $p$ | Identification rate | 0.01 -0.1 | (17) |

## 4. Results

This work defines the pandemic peak 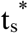 by the time Y attains its maximum in 1 year, that is 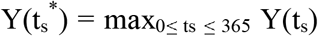. Firstly, we set the identification probability p = 0.1. In this case, we estimate the following predictions on the long time behavior of Y(t) for β = 0.35, 0.33, and 0.31.

We see from the estimated epidemic peak is ts* = 186(95%CI, 174-199). That is, starting from 1^st^ March 2020 (t_s_ =0), the estimated epidemic peak is 3^rd^ October 2020 (t_s_ =186), and the uncertainty range is from 22^nd^ August 2020 (t_s_ =174) to 16^th^ September 2020(t_s_ =199).

### 4.1. The possible effect of interventions

Interventions such as lockdown, quarantine, face masks, gloves reduce the number of cases reported more significantly. These measures can reduce the infection rate and will possibly delay the epidemic peak for approximately one month.

We have estimated the COVID-19 spread for a 40 days lockdown in India from (24^th^ March to 3^rd^ May) (36). We obtain the result given in Fig 3 as a result, when using the SEIR model for the identified parameters in Table 2. The estimated epidemic peak has been in the time slice 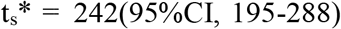 (95%CI, 195-288). The calculation has been made from the 1^st^ March (t_s_ =0), the estimated peak is 29^th^ October 2020(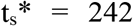), and the uncertainty peak day ranges from 12^th^ September(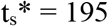) to 14^th^ December(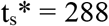).

**Fig 3.**
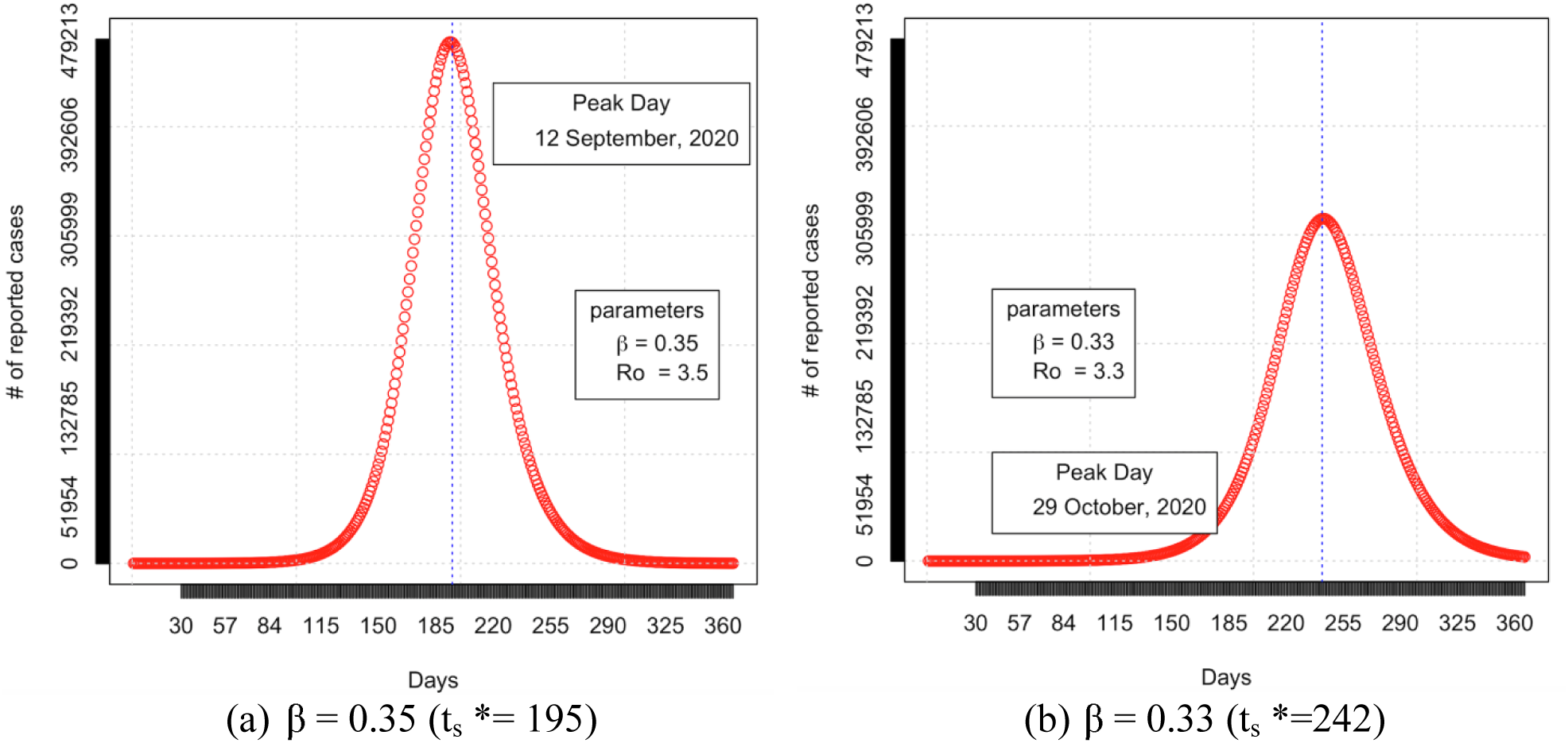
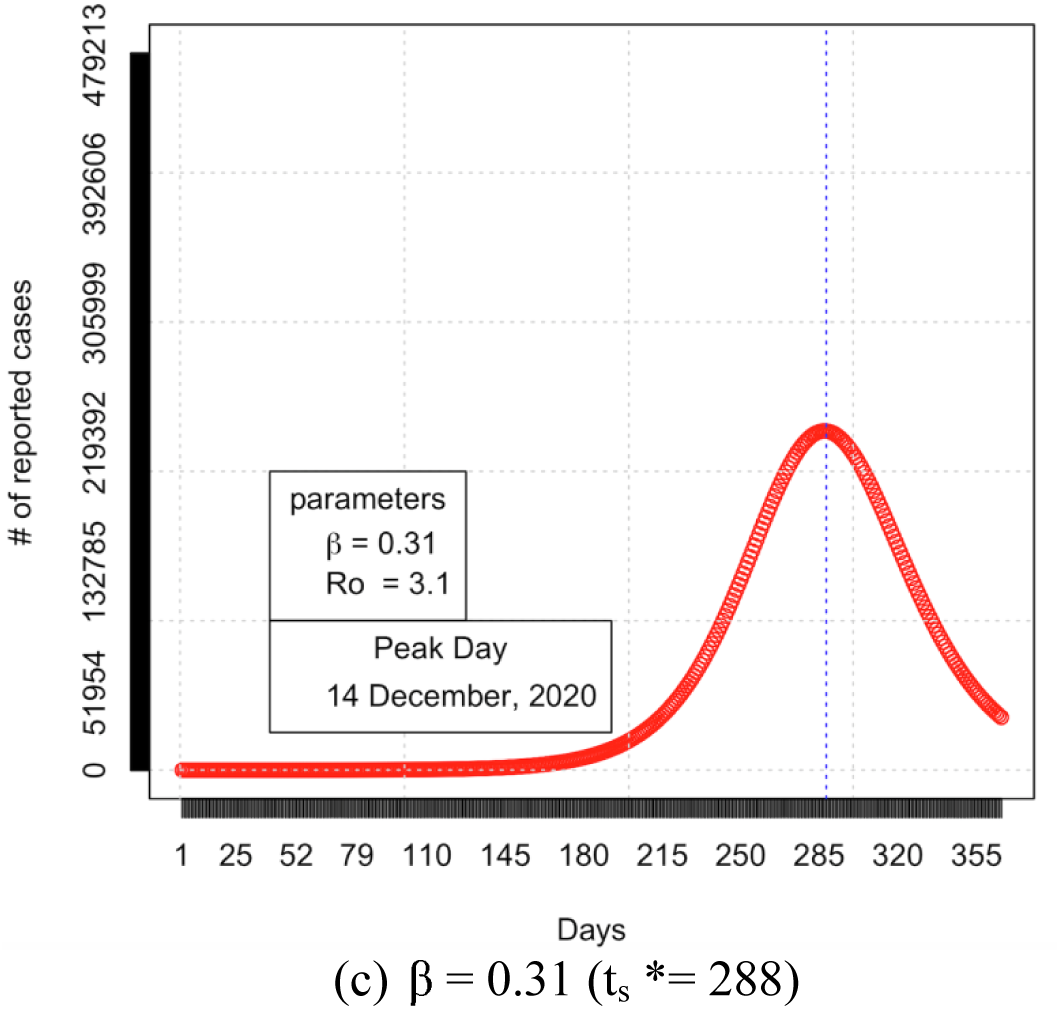
Time variation of the number Y(t_s_) of the infective individuals who are identified at the time slice t_s_ (0< t_s_ <365) for after 40 days of lockdown. The dotted lines represent the epidemic peak 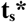.

After analyzing the SEIR model prediction, India’s planned lockdown period 40 days has been introduced to the model. The rate of spread has been reduced for those 40 days. Fig 4 clearly shows that the intervention has a positive effect on the delay of the epidemic peak. This delay in the epidemic peak can help in the improvement in the medical environment and research to treat the patients well. Fig 3 and Fig 4 show that there is a small change for a minimal period t_s_ <= 130. Therefore the interventions like government restrictions for vehicle movement and lockdown should be extended to reduce the final epidemic size.

The comparison of the predicted infected patients with and without lockdown has been depicted in Fig 5. It shows clearly that the lockdown has created a positive effect in reducing the risk of COVID-19 infection. Flattening the curve will reduce the overload in the health care system of India. India’s healthcare facilities are not sufficient (37) to handle any massive outbreak when considering the population size and very small doctors per 1000 citizens ratio; hence the country needs to depend on alternative strategies. Flattening the infection curve should also help to reduce the death rate in the country. From the history of Spanish flu during 1918 it is clearly evident that lifting the lockdown early may create the next peaks which may cause even more deaths than the first wave (38).

**Fig 5:**
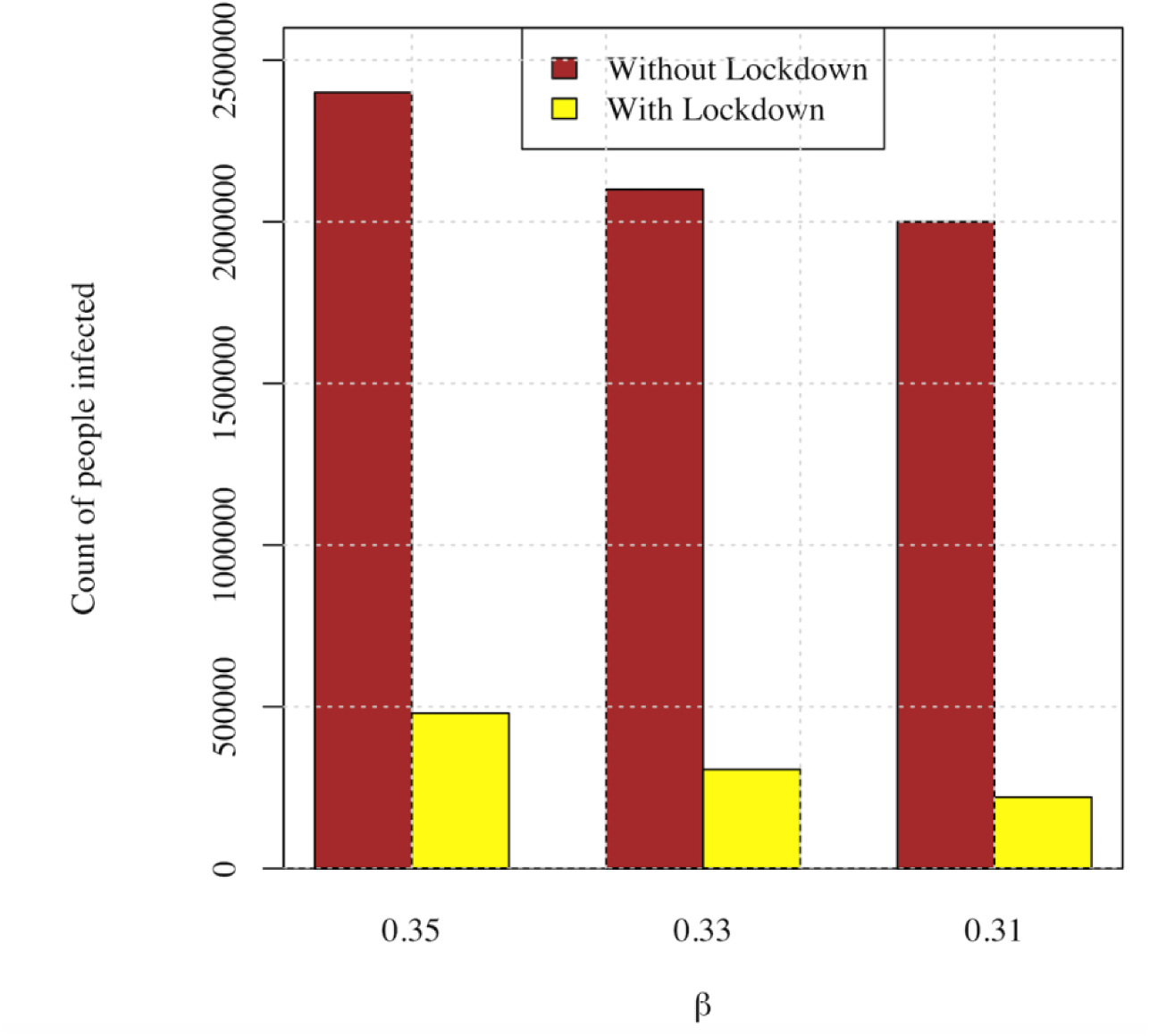
Comparision of the number of predicted infected cases without and with lockdown.

## 5. Discussion

As of May 5, 2020, the total number of COVID-19 infected and the death toll rose to 32,138 and 1,568, respectively. An increase in active cases is an alarming situation because, as discussed earlier, India is one of the largest population powerhouses in the whole world. Also, India’s healthcare infrastructure is week when we compare it with the WHO guidelines and one of the the worst doctor-people ratio (i.e., 1:11,082) in the world (The Financial Express, 2018). India will enter into STAGE III (40) of spreading if we do not correctly predict and implement the right methodologies to stop the spreading. In the present scenario, there is no proper vaccine or drugs for this deadly pandemic. Mathematical modeling like this research can help us predict the intensity of this pandemic and can effectively fix it with necessary measures. During this study, the major challenge was with the data. The data was not linear, and it showed an exponential growth concerning the infected cases in India. Over fitting has been another significant problem with a number of infected time series data.

In this study, we applied the SEIR compartmental model to the COVID-19 daily reported cases in India from March 1^st^, 2020, to April 14^th^, 2020. We have estimated the transmission rate as 0. 33(95%CI, 0.31-0.35) and the R_0_ the basic reproduction number as 3.3(95%CI, 3.1-3.5) and predicted the epidemic peak would be reached possibly in the September. Our prediction shows that India would have reached 2,400,000 infected people by the end of May 2020 if the lockdown was not pronounced. Since the government announced lockdown, India should expect the epidemic peak by September 2020 with lower number of cases which will be approximately around 450,000.

Our research also makes it clear that the epidemic COVID-19 in India is not going to end quickly. The estimated values of the R_0_ in this work is not different from the other papers (table 2). Finally, we provide a suggestion for the Indian Govt. and Policymakers to take the following steps:

- the intervention, like lockdown and other measures by the government, has a positive effect on delaying the epidemic peak, which will improve the medical environment and give many research results before the peak;
- intervention over a relatively long period is required to reduce the size of COVID-19 pandemic effectively;
- India needs to increase the number of tests to find the percentage of infected cases in different states and regions.

For this research, we have assumed the same transmission probability, removal rate, and the onset value throughout this work. These rates may increase or decrease when days passed by according to the intensity of the pandemic. The real-time change in daily data may change the predictions accordingly. According to some Indian researches, many other factors, such as “immune power in gene because of environment, and food habits,” can help India to flatten the infection curve (41,42).

## Data Availability

All data used in the manuscript has been taken from the Internet sources.
https://www.mohfw.gov.in/
https://www.worldometers.info/coronavirus/
https://www.who.int/emergencies/diseases/novel-coronavirus-2019

## Acknowledgement

This work was partially supported by Polish National Science Centre, decisions no. 2016/21/D/ST6/02408 and Wrocław University of Science and Technology statutory funds.

